# Telomere length and mitochondrial copy number as potential biomarkers for male infertility in Iraqi men

**DOI:** 10.1101/2025.05.11.25327013

**Authors:** Mustafa Faeq Kadhim, Mehdi El Arbi, Farah Thamer Samawi, Ali Gargouri

**Author notes:** Corresponding author: Ali Gargouri.

## Abstract

Male infertility is one of the major health and social problems in Iraq. It can be caused by several factors, such as acquired, environmental, and genetic factors. Awareness of the crucial role of telomeres and mitochondria in sperm production and fertility has increased in recent years. This study aims to evaluate the relationship between mitochondrial DNA (mtDNA) copy number and telomere length in sperm and the degree of infertility in Iraqi men. Of the 200 study participants, 50 were healthy controls and 150 were infertile. Sperm count, motility, and morphology were assessed by collecting and analyzing semen samples. The mitochondrial gene ND1 and the nuclear reference gene GAPDH were analyzed by quantitative PCR (qPCR) to determine the mtDNA copy number, while another qPCR analysis was used to determine telomere length. The mtDNA copy number of infertile men was significantly higher than that of healthy controls, with a p-value of 0.001. In addition, sperm from the patient group showed a significant reduction in telomere length (p = 0.001). According to the study results, male infertility in Iraqi men is associated with a higher mtDNA copy number and shorter telomere length. DNA damage or disruption of the mitochondrial energy production pathway could be the cause of this association. These results highlight the importance of conducting further investigations to validate these findings and understand the underlying biological pathways, as well as to open new research perspectives in the field of male infertility.

**Highlights:**

- Infertile men have altered mtDNA copy number compared to healthy ones.
- Telomere length is a biomarker of aging and reproductive health.
- Oxidative stress and impaired spermatogenesis are thought to contribute to telomere shortening.
- Shorter telomere length is associated with decreased sperm quality and fertility.

## Introduction

Infertility is a major social and psychological problem for couples in Iraq. It is defined as the inability to conceive after one year of unprotected sexual activity for women under 35 or six months for those over 35[1]. Female factors account for about one third of infertility cases, followed by male factors for another third, and a combination of factors or unexplained reasons for the remainder [2]. Infertility can be attributed to a range of risk factors, including advanced age, smoking, alcoholism, obesity, malnutrition, and sexually transmitted infections. Global estimates of the prevalence of infertility range from 3.5% to 16.7% in developed countries and from 6.9% to 9.3% in underdeveloped countries[3,4].

In Iraq, infertility has a major effect on the sexual activity, self-image, and self-esteem of both men and women. Infertility has become much more common in Iraq over the past 16 years (2000–2016). The consequences of war, daily stress, smoking, work-related hazards, dietary habits, and genetic predispositions represent some of the causes associated with this trend [5].

While childbirth is often seen as a fundamental part of female identity, being a parent is seen by many Iraqi men as a sign of adulthood[5]. The high prevalence of infertility in Iraq is illustrated by the growth of IVF centers and infertility clinics in the country. Infertility in Iraq is poorly understood, despite its importance. The growing body of research on infertility serves as a bridge to fill the knowledge gap and improve our understanding of infertility and its negative impact on Iraqi society [6].

The World Health Organization (WHO) defines primary infertility as the inability of a couple to conceive despite regular, unprotected sexual intercourse. Individuals who have never been pregnant are influenced by this condition. Secondary Infertility occurs when a couple who has previously conceived fails to conceive again within a year of unprotected sexual activity. More than 80 million people worldwide suffer from infertility, affecting 10–15% of couples. Secondary infertility is more common than primary infertility [7].

The amount of mitochondrial DNA (mtDNA) molecules present in a cell is indicated by its copy number. The "powerhouses of the cell," or mitochondria, are organelles that produce energy. They contain DNA that is different from the nuclear DNA that exists in the nucleus of the cell. Mitochondria play a critical role in sperm health by producing the energy needed for sperm production and function. A reduced number of mtDNA copies can compromise this energy production, leading to impaired sperm motility, viability, and overall fertility[8,9].

Male fertility is strongly influenced by mitochondrial DNA copy number. Reduced energy production, oxidative stress, apoptosis, and poor sperm maturation are all consequences of increased copy number that can lead to infertility. A better understanding of the link between male infertility and mtDNA copy number could help in the creation of new diagnostic and therapeutic approaches[10]. The current study was conducted to evaluate mtDNA copy number and telomere length in infertile men, compared to a control group.

## Materials and methods

### Subjects

This study included 200 semen samples from 150 infertile Iraqi men and 50 fertile Iraqi men (used as a control group). The infertile men were classified into three groups based on their semen parameters:

- **Group 1 (A):** 49 men with asthenozoospermia
- **Group 2 (OA):** 44 men with oligoasthenozoospermia
- **Group 3 (OAT):** 57 men with oligoasthenoteratozoospermia

### Ethical Approval

Semen samples were collected from the High Institute for Infertility Diagnosis and Assisted Reproductive Technologies 12 April 2023 and 5 February 2024. The study, conducted at Al-Nahrain University’s Forensic DNA Research and Training Center, adhered to the ethical principles outlined in the Declaration of Helsinki. All participants provided informed written consent before sample collection.

### Semen Analysis

#### Macroscopic Examination

Appearance: Typically opalescent or grayish-white. Abnormal colors, such as brown or red, may indicate the presence of blood. Volume: measured using a graduated test tube. The normal volume varies from 1.5 to 5.0 milliliter per ejaculation. Hypovolemia is diagnosed when the volume is less than 1.5 ml. pH: Measured using Litmus paper. The normal range is 7-8.0. Liquefaction: should occur within 30 minutes to 1 hour at 37°C or room temperature. Viscosity: assessed by observing the flow of semen through a pipette. Normal semen should form a single small drop and flow smoothly.

#### Microscopic Examination

Sperm Concentration: determined by counting sperm in a small sample under a microscope and multiplying by a factor to estimate the concentration per milliliter. The normal sperm count varies from 20 to 150 million sperm per milliliter. Sperm Motility: evaluated by observing the movement of sperm under a microscope. Motility is categorized as progressive, non-progressive, or immotile. Sperm Morphology: assessed by examining the shape and structure of sperm under a microscope. Normal sperm morphology is characterized by a specific shape and size.

#### Sperm DNA Extraction, Purity and Concentration

According to the manufacturer’s instructions, DNA is extracted from sperm using a spin column chromatography technique with the Presto™ Sperm DNA Extraction Kit (cat.no. GSP100), manufactured by Geneaid, Taiwan. By lysing the sperm cells and degrading proteins, the process allows the DNA to attach to the transparent fiber matrix of the spin column. A low-salt elution buffer is then used to elute the pure DNA after removing contaminants using wash buffers. No alcohol precipitation or phenol/chloroform extraction is required, and the entire process can be completed in just ten minutes. PCR and other enzymatic reactions can be performed with the purified DNA. DNA concentration and purity were assessed using aOneCNanodrop spectrophotometer (Thermo Fisher Scientific, USA). DNA purity was determined by calculating the A260/A280 ratio, which should ideally be between 1.7 and 1.9. A ratio within this range indicates a pure DNA sample.

### Quantification of Sperm mtDNA Copy Number by Real-time PCR

#### Principle

The most common methods to measure mtDNA copies are PCR-based, with qPCR being the most widely used due to its simplicity. Most protocols assess relative mtDNA copy number by comparing mtDNA levels to a nuclear gene, making comparisons between studies difficult. This study followed this procedure to ensure the accuracy of the results and obtain valuable feedback. Absolute mtDNA copy number can be determined using qPCR with a standard curve[11,12]. In the current study, the mitochondrial gene NADH dehydrogenase subunit 1 (ND1) served as the target for mtDNA quantification while Glyceraldehyde-3-phosphate dehydrogenase (GAPDH) was used as the reference nuclear gene.

#### Procedure

Relative mitochondrial DNA (mtDNA) content was quantified using real-time polymerase chain reaction (PCR) targeting the ND1 gene and normalized by simultaneously measuring the nuclear DNA (nDNA)-encoded GAPDH gene. qPCR was performed using a Real-Time quantitative PCR System (Qiagen, Germany) and used a specific primer set showed in Table 1. A reaction mixture was prepared separately for the mtDNA ND1 gene and GAPDH with a final volume of 20 µL in different tubes. Each reaction contained 10 µL of TransStart® Tip Green qPCRSuperMix (TransGen, biotech. AQ141-01) containing the fluorescent dye Eva green, 1 µL of forward and reverse primers and 4 µL of DNA sample (diluted with TBE buffer and measured by nanodrop at 260/280 nm wavelength) for both mtDNA and nDNA. A non-template control (NTC) was prepared containing all components of the master mix except template DNA. The thermal profile of the quantitative PCR used to estimate copy number is described as follows: 94°C for 1 min, followed by 35 cycles (Denaturation at 94°C for 10 sec, Annealing 58°C for 20 sec, Extension 72°C for 1 min). A melting temperature range of 55°C to 95°C was adopted to perform the melting curve and to confirm that no primer dimers were found.

**Table 1:**
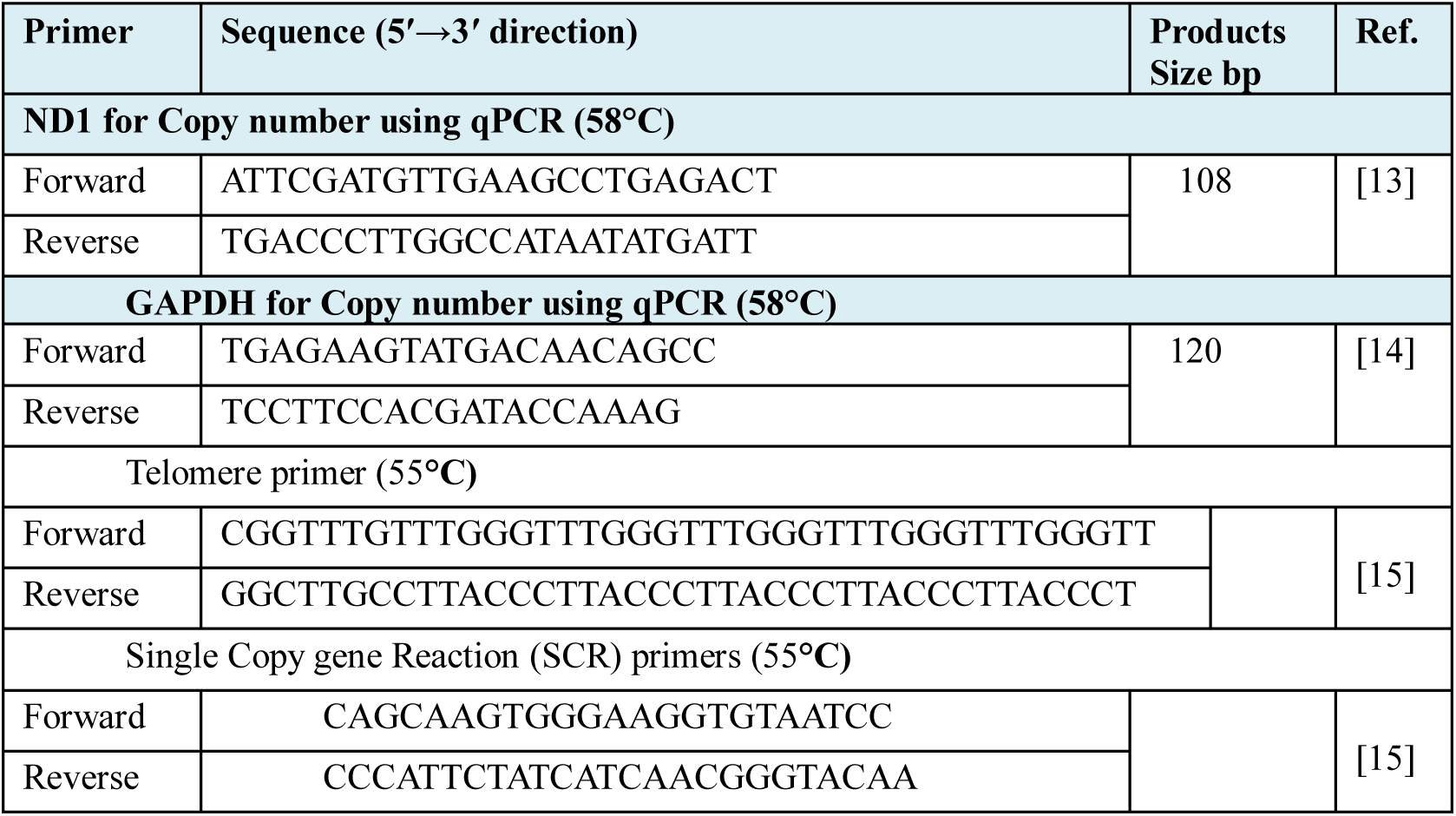
The primer sequence used in the current study.

### Quantification of Sperm Telomere Length (STL) expression by qPCR

#### Principles

Human Telomere Length Quantification by qPCR Assay Kit(Cat.no. EQ022-01) manufactured by (ELK Biotechnology company, Wuhan, China). The single copy gene (SCR for Single Copy gene Reaction) primers are designed to specifically recognize and amplify a 78 bp region of human chromosome 11. Quantitative Polymerase Chain Reaction (qPCR) is a powerful technique for measuring the relative abundance of specific DNA sequences. This method was adapted to quantify sperm telomere length. Relative Quantification using the 2-ΔΔCT method was used. The 2-ΔΔCT method compares the cycle threshold (CT) values of a target gene (telomere repeat sequence) to a reference gene (single-copy gene, known as36B4, Large ribosomal subunit protein uL10) which encodes the acidic ribosomal phoshoprotein P0 (RPLP0).Table 1 showed the primer sequence for SCR and TP genes The difference between the CT values (ΔCT) represents the relative abundance of the target gene normalized to the reference gene. A larger ΔCT indicates a lower abundance of the target gene. Finally, accurate ΔΔCT calculations require approximately equal amplification efficiencies for target and reference genes. This must be validated for each specific study[16].

#### Procedure and Reaction Components

The EnTurbo™ SYBR Green PCR SuperMix provides specific and sensitive detection over a wide range of DNA concentrations. The SYBR Green I dye binds to double-stranded DNA, allowing non-specific detection of amplified products while the Hot-Start Taq Polymerase prevents extension of primersbinding to templatesequenceswithlowhomology (mispriming) as well as the extension of primersbinding to eachother (primer-dimer formation) duringreaction setup. The PCR reaction is optimized for low-concentration templates to ensure accurate quantification. For each sample, two separate 20 µL PCR reactions were set up to independently amplify the single-copy gene and the telomere repeat (TP) regions. The specific reagents used for these reactions included (0.4µL) primer stock solution (Telomere or SCR), 4µL Genomic DNA Template, Reaction mix (10µL) and (5.6µL) RNase-free ddH2O. Amplification was performed using a thermal cycler programmed with the temperature profile described as (Initial denaturation at 95°C for 1 min, 40 cycles (Denaturation at 95°C for 30 sec, Annealing at 55°C for 30 sec, Extension at 72°C for 1 min) and the same melting temperature range (55°C to 95°C) for melt curve profiling.

### Statistical Analysis

IBM Statistical Package for the Social Sciences (SPSS) version (26) and GraphPad Prism version (8) were used for descriptive and numerical statistics [17]. Using qPCR to quantify the relative abundance of mtDNA, by comparing the expression of the target gene (ND1) to a reference gene (GAPDH), the relative mtDNA content is determined. Telomere length results were analyzed according to theΔCT and ΔΔCT equations performed as in[18,19].

## Results

### Age of the study Groups

The mean age ± SE for the patient group was (33.047± 0.81) while in the control group it was (30.880 ± 1.43). The results showed that there was no significant difference in age between the two groups (P=0.1).

### Demographic characteristics

Table 3 shows that, compared with the control group, a significantly higher percentage of patients had a family history of the disease (p < 0.001). The study results showed that there were notable variations between patient and control groups in a number of biological, behavioral, and demographic factors that contributed to infertility. We show that smoking, education level, length of infertility, and family history of fertility are important contributors in male infertility, table 3.

**Table 2:** Age comparsion among study group.

| Parameters | Group | Mean | Std. Error<br>Mean | P-value |
| --- | --- | --- | --- | --- |
| Age | Patients | 33.047 | 0.81 | 0.1 N.S |
|  | Control | 30.880 | 1.43 |  |

**Table 3:** Dempgraphic variable in patients and control group.

| Parameters |  | Group |  | Chi-square | P-value |
| --- | --- | --- | --- | --- | --- |
|  |  | Patients | Control |  |  |
| Family history | Yes | 67 (44.67%) | 1 (2%) | 30.42 | 0.001** |
|  | No | 83 (55.33%) | 49 (98%) |  |  |
| Alcohol | Yes | 66 (44%) | 6 (12%) | 16.67 | 0.001** |
|  | No | 84 (56%) | 44 (88%) |  |  |
| Smoking | Yes | 48 (32%) | 21 (42%) | 1.66 | 0.2 |
|  | No | 102 (68%) | 29 (58%) |  |  |
| Treatment | Yes | 102 (68%) | 12 (24%) | 29.62 | 0.001** |
|  | No | 48 (32%) | 38 (76%) |  |  |
| Wife miscarriage | Yes | 25 (16.67%) | 0 | 9.52 | 0.002 |
|  | No | 125 (83.33%) | 50 (100%) |  |  |
| Education Level | No. Edu | 62 (41.33%) | 7 (14%) | 84.46 | 0.001** |
|  | P.S | 11 (7.33%) | 0 |  |  |
|  | S.S | 57(38%) | 2 (4%) |  |  |
|  | University | 20 (13.33%) | 41 (82%) |  |  |
| Infertility<br>Duration | 1-5 Years | 74 (49.33%) | 0 | 200.00 | 0.001** |
|  | 6-10<br>Years | 32 (21.33%) | 0 |  |  |
|  | >10 years | 44(29.33%) | 0 |  |  |
|  | No | 0 | 50 |  |  |

\* No. Education, P.S Primary School, S.S secondary school

These results suggest that the onset of the disease could have a genetic or environmental influence. Although the patient group has a higher percentage of alcohol consumers, the difference is not statistically significant (p = 0.01). This demonstrates that alcohol consumption, although it may be a risk factor, may not be an important indicator of the disease. There is also no statistically significant difference in the prevalence of smoking between the two groups (p value = 0.2). As with alcohol consumption, it also appears that smoking is not a significant risk factor for this particular disorder.

Treatment protocol may influence the outcome of the disease, as shown by the significant difference in treatment rates (p < 0.001). Further research on the effectiveness of different treatment approaches is needed. The percentage of miscarriages in wives of patients was significantly higher (p < 0.001). This can be a sign of underlying reproductive health problems or other variables that influence the disease and miscarriage. Given the significant difference in education levels (p < 0.001), it is possible that a higher risk of the disease may be related to lower education levels. Several variables, such as economic status, access to health care or lifestyle choices, could be to responsible for infertility. Longer periods of infertility may be attributed to an increased risk of the disease or may be the result of increased complication of infertility, as indicated by the substantial difference in duration of infertility (p < 0.001). According to the table 3, infertility may be the result of a confluence of environmental, lifestyle, and genetic variables. The condition appeares to be significantly associated with factors such as family history, education level, and reproductive health problems. To clarify the underlying mechanisms and create efficient prevention and treatment strategies, further research is needed.

### Seminal fluid parameters

In terms of sample volume and sperm count, the study found statistically significant differences between the patient group and the control group, table 4. The mean sample volume of the patient group was significantly smaller than that of the control group (3.11 mL versus 4.59 mL, p < 0.001). The patient group also had a significantly reduced mean sperm count (15,947 versus 39,800 millions/ml sperm, p < 0.001). These findings suggest a potential link between reduced sample volume and sperm count and the medical condition studied. Both of these variables were significantly lower in the patient group than in the control group. These results could have important therapeutic implications and need to be confirmed by further research.

**Table 4:** Asample volume and sperm count in patients and control group.

| Parameters | Group | Mean | Std. Error<br>Mean | P-value |
| --- | --- | --- | --- | --- |
| Volume (ml) | Patients | 3.110 | 0.14 | 0.001** |
|  | Control | 4.590 | 0.12 |  |
| Sperm count<br>(million/ ml) | Patients | 15.947 | 1.59 | 0.001** |
|  | Control | 39.800 | 2.44 |  |
|  | **Character** | **Patients** | **Control** |  |
|  | Normal | 100 (66.67%) | 50 | 0.001** |
| Morphology of Sperm | Abnormal | 50 (33.33%) | 0 |  |
| Sperm Motility | PR | 89 (59.33%) | 50 | 0.001** |
|  | NP | 30 (20%) | 0 |  |
|  | IM | 31 (20.67%) | 0 |  |

Male infertility variables may contribute to the etiology of the disease, as evidenced by significant variations in sperm motility and morphology (p < 0.001).

### mtDNA copy number and telomere length

Telomere length and mtDNA copy number are two variables that are statistically compared between several patient groups and a control group in the data tables presented. The mean values of the two variables are compared in Figure 1 below and (Table I in supplementary file) between the control group and the patient group. The standard error of the mean is also shown, which helps to measure the precision of the mean. The difference between the two groups is statistically significant, as indicated by the p-value. The result showed that Patients have a significantly higher mean number of mtDNA copy than controls (p < 0.001). In terms of Telomere Length, Patients had a considerably lower mean telomere length than controls (p < 0.001).

**Figure 1:**
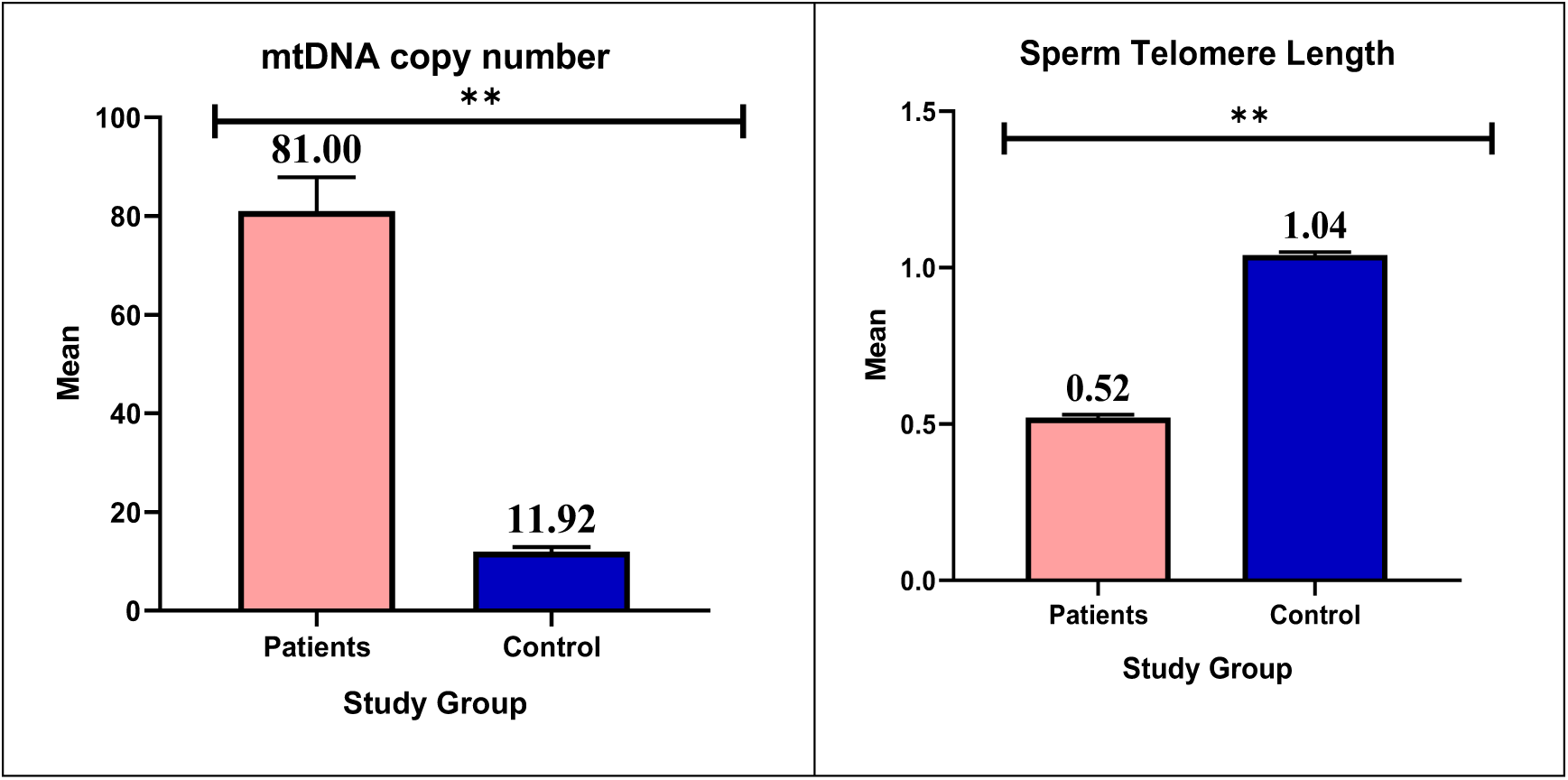
Mean and Standard Error of mtDNA copy number and telomere length in patients and control

The qPCR result of the ND1 gene is shown as an example in Supplementary Figure I. Supplementary Figure II shows the qRT-PCR result of telomere length. Mean values of mtDNA copy number and telomere length can be distinguished in Figure 2 (supplementary Table II) between the control group and various subgroups of patients. Significant differences between goups are indicated by the letters a, b, and c. Patients with asthenozoospermia have significantlylower copy number than those in the control, oligoasthenozoospermia, and oligoasthenoteratozoospermia. Compared with the oligoasthenoteratozoospermia and control groups, oligoasthenozoospermia patients have significantlyreduced copy numbers. Patients with oligoasthenoteratozoospermia have significantlyhigher copy numbers than any other group.

**Figure 2:**
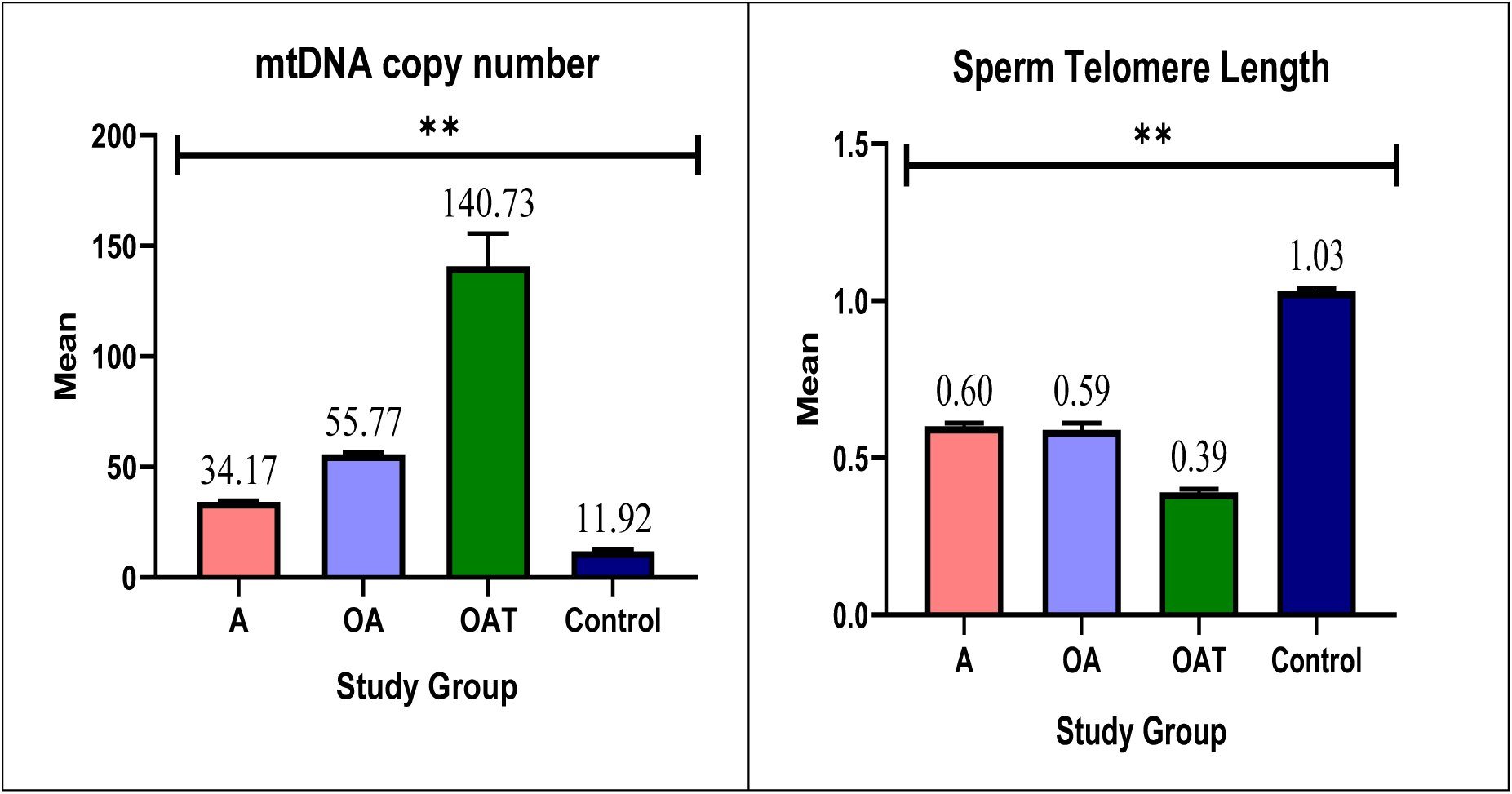
Comparison in copy number and telomere length in patients subgroups and control group; **A:** Asthenozoospermia, **OA:** Oligoasthenozoospermia, **OAT:**Oligoasthenoteratozoospermia

Telomere length is significantly reduced in patients with asthenozoospermia and oligoasthenozoospermia compared withthe control group. Patients with oligoasthenoteratozoospermia express the least amount of telomere length among all other groups. The results imply that, compared with healthy controls, patients with different levels of male infertility have notable changes in both copy number and telomere length. These results could be used as biomarkers for diagnosis and prognosis and may shed light on the underlying causes of male infertility.

Patients with Oligoasthenoteratozoospermia had a significantly higher copy number than any other group, including the control group. This implies that this specific subgroup has abnormal gene copying machinery. Compared with all other groups, patients with Asthenozoospermia showed a significantly lower copy number. Moreover, compared to with both the Oligoasthenoteratozoospermia and control groups, patients with Oligoasthenozoospermia had a milder reduction. This suggests that within these two groups (Oligoasthenoteratozoospermia andOligoasthenozoospermia), there is a mutation or defect in specific genes responsible for maintaining the number of mtDNA copies within a normal range. Regardless of all patient groups, the control group had the lowest copy number value, indicating that changes in copy number could indicate fertility complication.

Overall, the mean telomere length of the patient subgroups was significantly shorter than that of the control group, which included healthy individuals. Telomere destruction, which is often related to aging and cellular damage, is indicated by this shortening in individuals. All patient groups showed an overall decrease in telomere length, but these groups differed statistically significantly from each other. Compared with all other groups, including Asthenozoospermia and Oligoasthenoteratozoospermia, the Oligoasthenoteratozoospermia group had the shortest mean telomere length. This implies that the reproductive cells in this group have more cellular damage. Finally, the telomere length of the Asthenozoospermia and Oligoasthenozoospermia groups was significantly shorter than that of the control group, suggesting that these groups had cellular DNA damage.

### Correlation test between mtDNA copy number and telomere length

According to the current study, there is a moderate negative correlation between the telomere length level and mtDNA copy number in all study groups, including patients subgroups and the control group. Both variables have a negative association, as indicated by the value of the correlation coefficient (r = -0.5565) which means that telomere length decreases when mtDNA copy number increase and vice versa. A reverse connection is represented by a negative value, while a strong negative association is indicated by a number around -1.

According to the coefficient of determination value (r^2^ = 0.3097), mtDNAcopy number variations account for 30.97% of the telomere length variation. This demonstrates that telomere length is modulated by several factors other than the mtDNA copy number. P-value (0.001) indicates a statistically significant correlation between the variables, indicating a substantial negative assosiation between mtDNA copy number and telomere length.Figure 3 illustrates the result. Supplementary Figures III, IV and V show the correlation tests between the different subgroups of patients with controls(Asthenozoospermia& Control) (Oligoasthenozoospermia& Control) and (Oligoasthenoteratozoospermia& Control).

**Figure 3:**
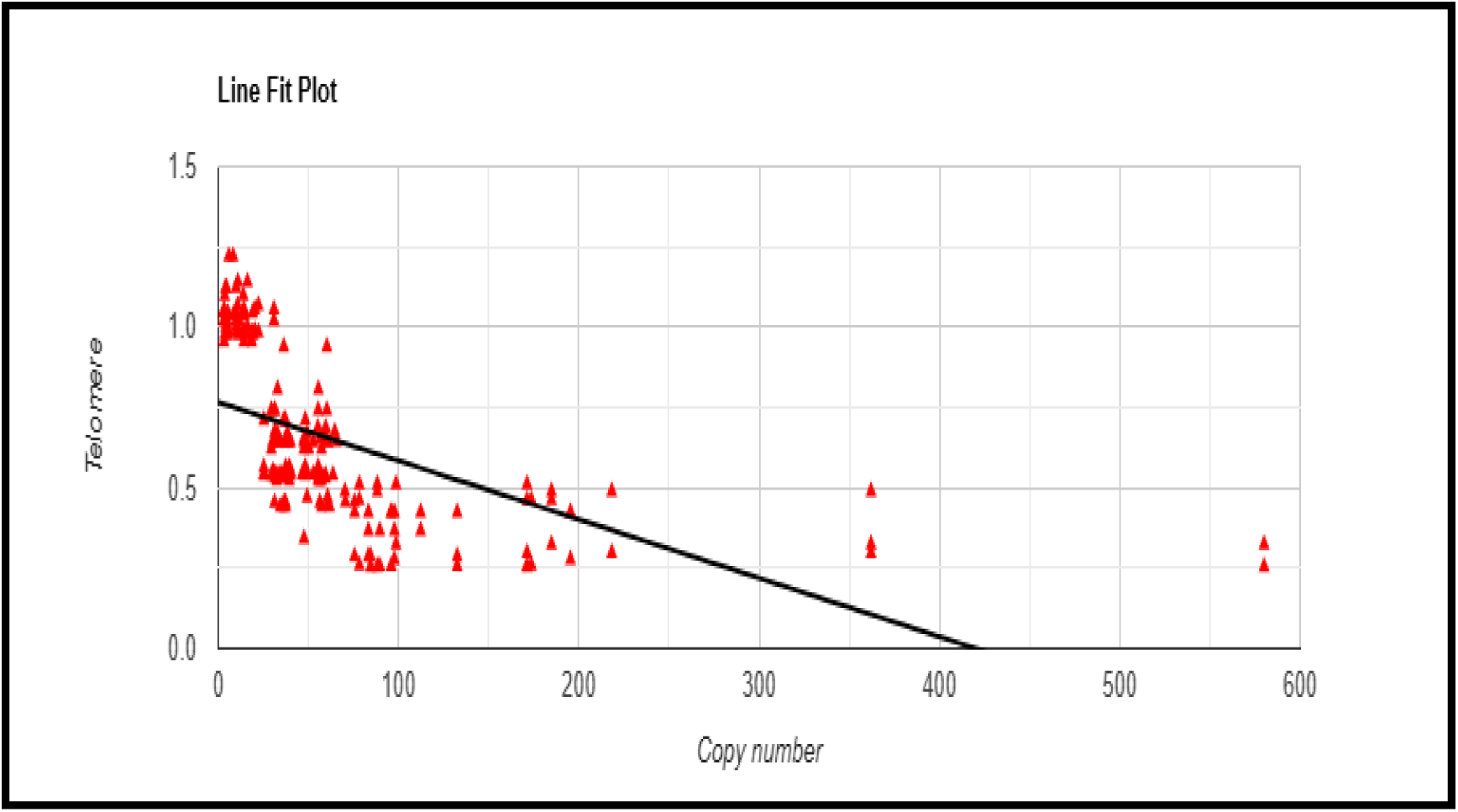
Correlation test between mtDNA copy number mean and telomere length level mean in all study group including patients subgroups and control

Correlation analysis between mtDNA copy number and STL (sperm telomere length) in patients with asthenozoospermia, oligoasthenozoospermia, and oligoasthenoteratozoospermia revealed an r^2^ value of 0.25 (r= -0.502), indicating that 25% of the variation in telomere length can be explained by changes in mtDNA copy number. However, the *p*-value˂0.01**, suggesting that this correlation is statistically significant This implies that although there is a weak to moderate relationship between these two variables, the association lacks strong statistical support, and the observed correlation may be influenced by other factors or sample variability (Figure 4).

**Figure 4:**
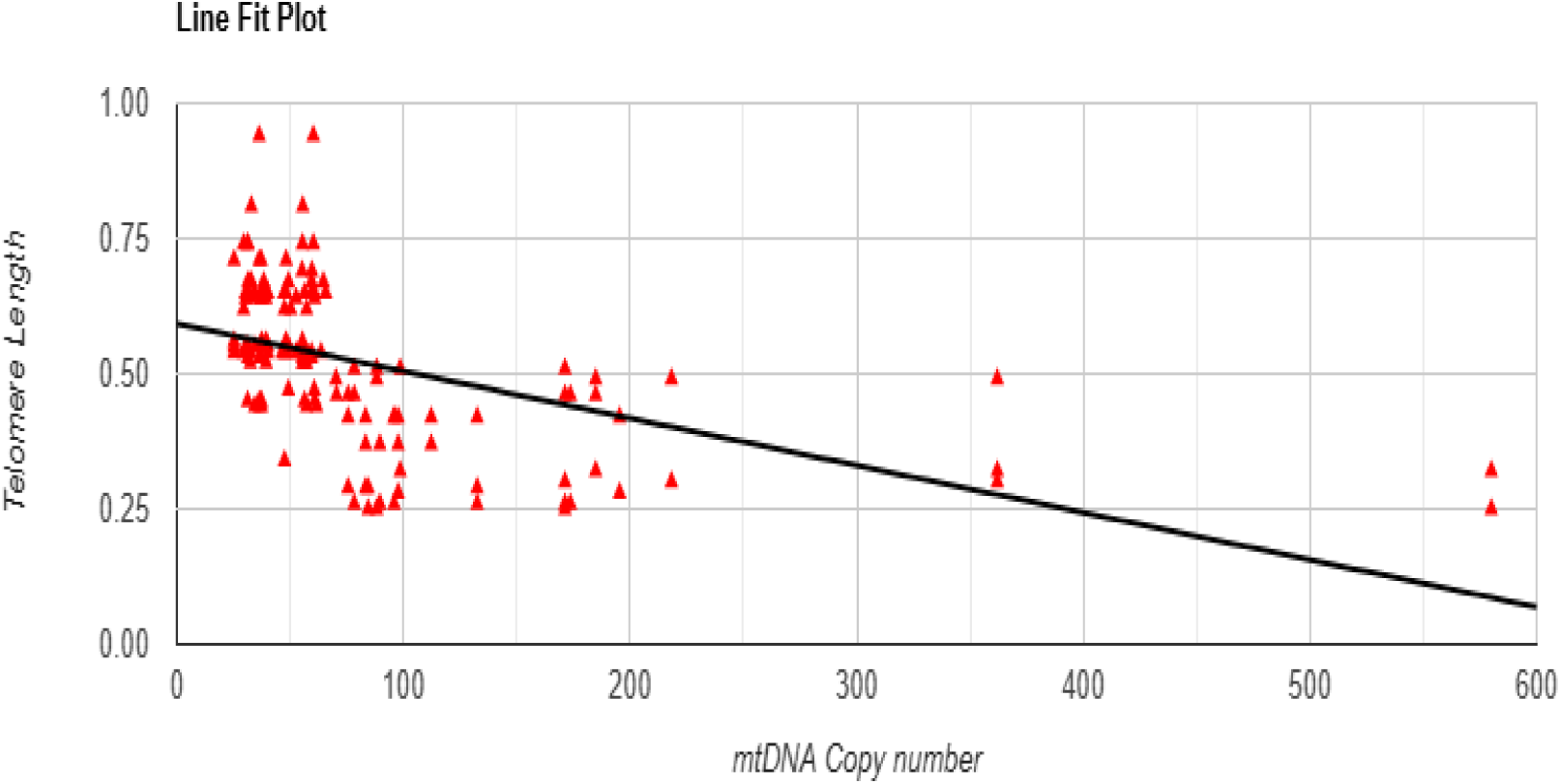
Correlation test between mtDNA copy number and telomere length in patients subgroups (asthenozoospermia, oligoasthenozoospermia and oligoasthenoteratozoospermia)with r^2^= 0.25 and p-value <0.01**.

Correlation analysis between mtDNA copy number and STL in the control group revealed an r^2^ value of 0.26, indicating that 26% of the variation in telomere length can be explained by changes in mtDNA copy number. However, the *p*-value was 0.2, which is not statistically significant (NS). This implies that even if there is a slight relationship, it is not strong enough to confirm a significant association, and other biological or methodological factors, as well as the increase in the number of control individuals, may influence the results.

### Linear regression test between mtDNA copy number and telomere length

Overall, a statistically significant linear association between mtDNA copy number and telomere length is provided by the regression model for all study group (Patients and control); the slope of the regression line is β = -0.0018, p <0.001 as showed in figure VI in supplementray file. This indicates that the expected telomere length decreases by an average of 0.0018 units for every unit increase in mtDNA copy number. As mentioned earlier in the correlation analysis, this negative coefficient denotes a negative association between the two variables. This coefficient is considered statistically significant if the p-value is less than 0.001. p <0.001, α = 0.77: This represents the intercept of the regression line. When the mtDNA copy number is zero, this indicates the expected telomere length. However, since a zero copy number may not be a realistic result, this interpretation may not be meaningful in the context of this particular study. The statistical significance of this intercept is indicated by the p-value <0.001.

### Receiver Operating Characteristic (ROC) curve analysis

The results of a Receiver Operating Characteristic (ROC) curve study are presented in Table 6. This study is frequently used to evaluate the performance of a diagnostic algorithm. In this case, the model uses two parameters: copy number and telomere length. The ability of the algorithm to distinguishbetween the two factors can be assessed by the area under the curve (AUC); values closer to 1 indicate greater accuracy. The threshold used to determine whether a sample is positive or negative is commonly referred to as the cutoff value. Specifically, specificity is the percentage of true negatives correctly identified, while sensitivity is the percentage of true positives correctly detected. The statistical significance of the results is illustrated by the p-value. Overall, the results suggest that mtDNA copy number and telomere length are good predictors, with copy number performing slightly better in terms of specificity. The AUC value was 0.954 and 0.89 for copy number and telomere length respectively, which is statistically significant (P< 0.001), while the cutoff value was 25.24 and 0.95 rcepectively. Table 5 and Figure 6 illustrated the results.

**Figure 5:**
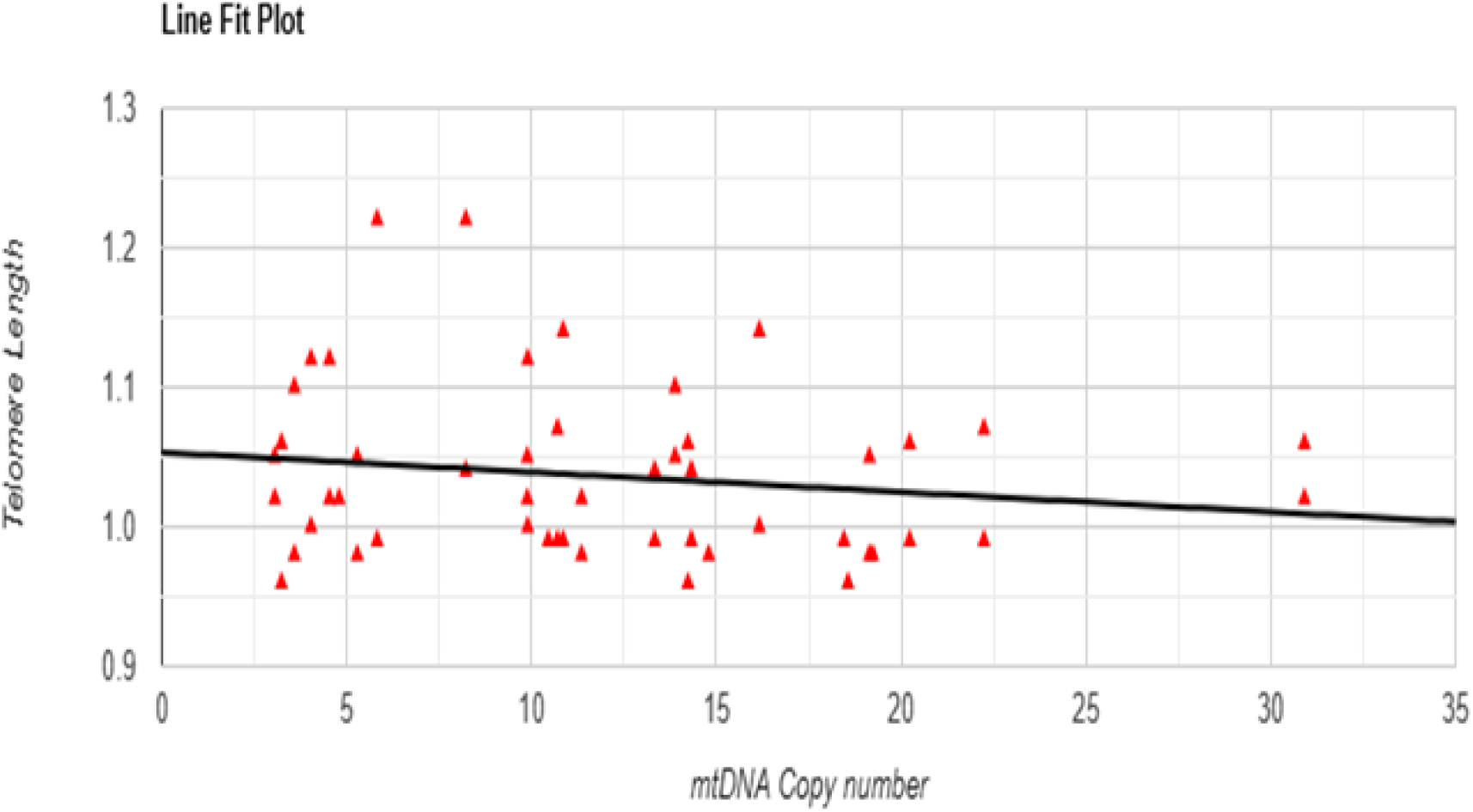
Correlation test between mtDNA copy number and telomere length in control group with r^2^= 0.26 and p-value =0.2 NS.

**Figure 6:**
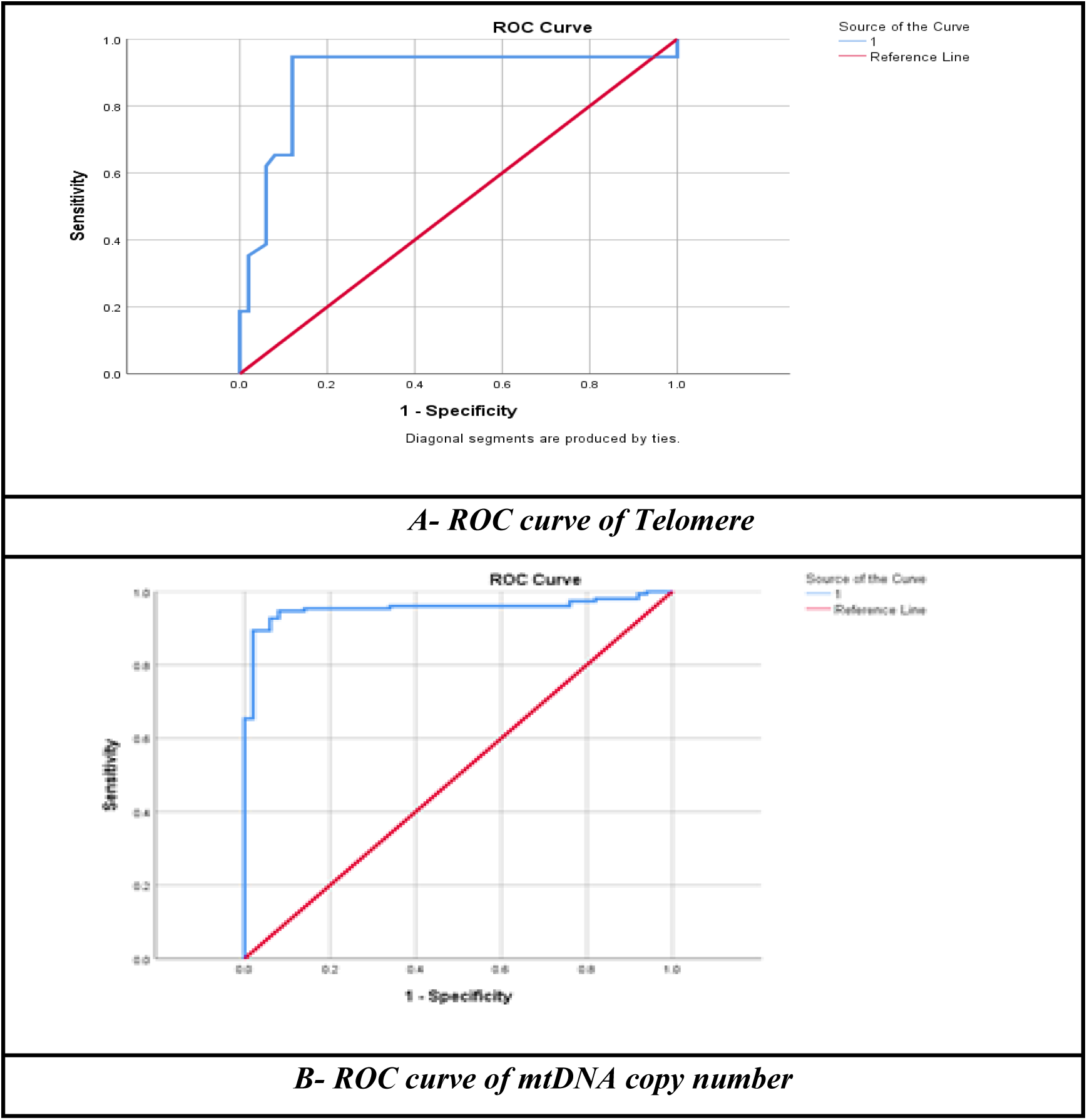
Receiver Operating Characteristic (ROC) curve of Sperm Telomere Length and mtDNA copy number.

**Table 5:**
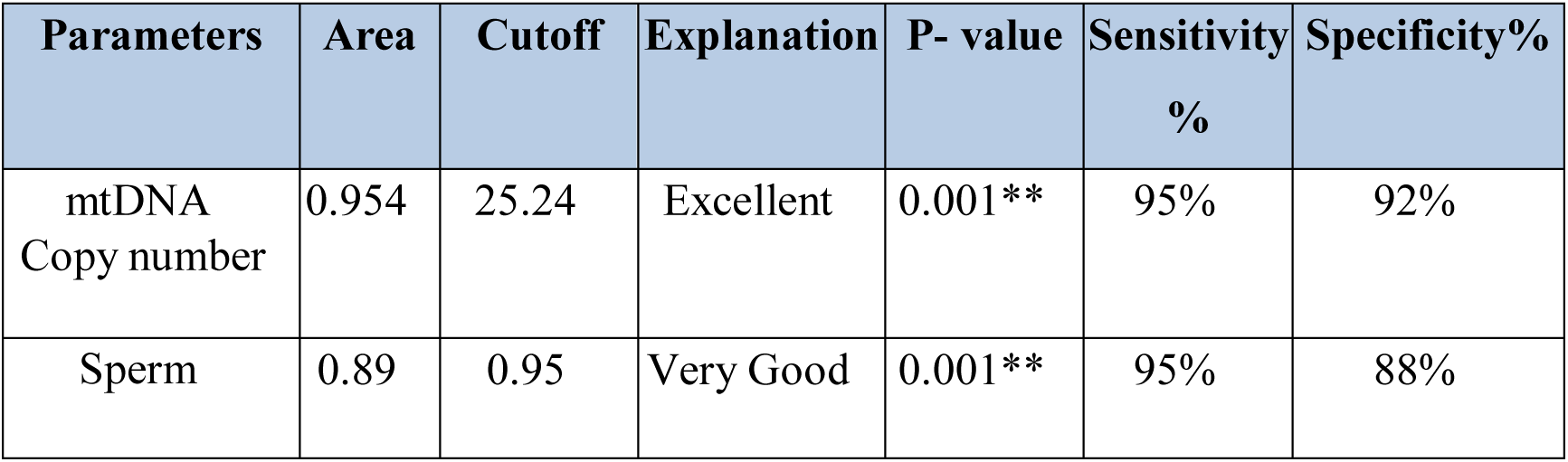
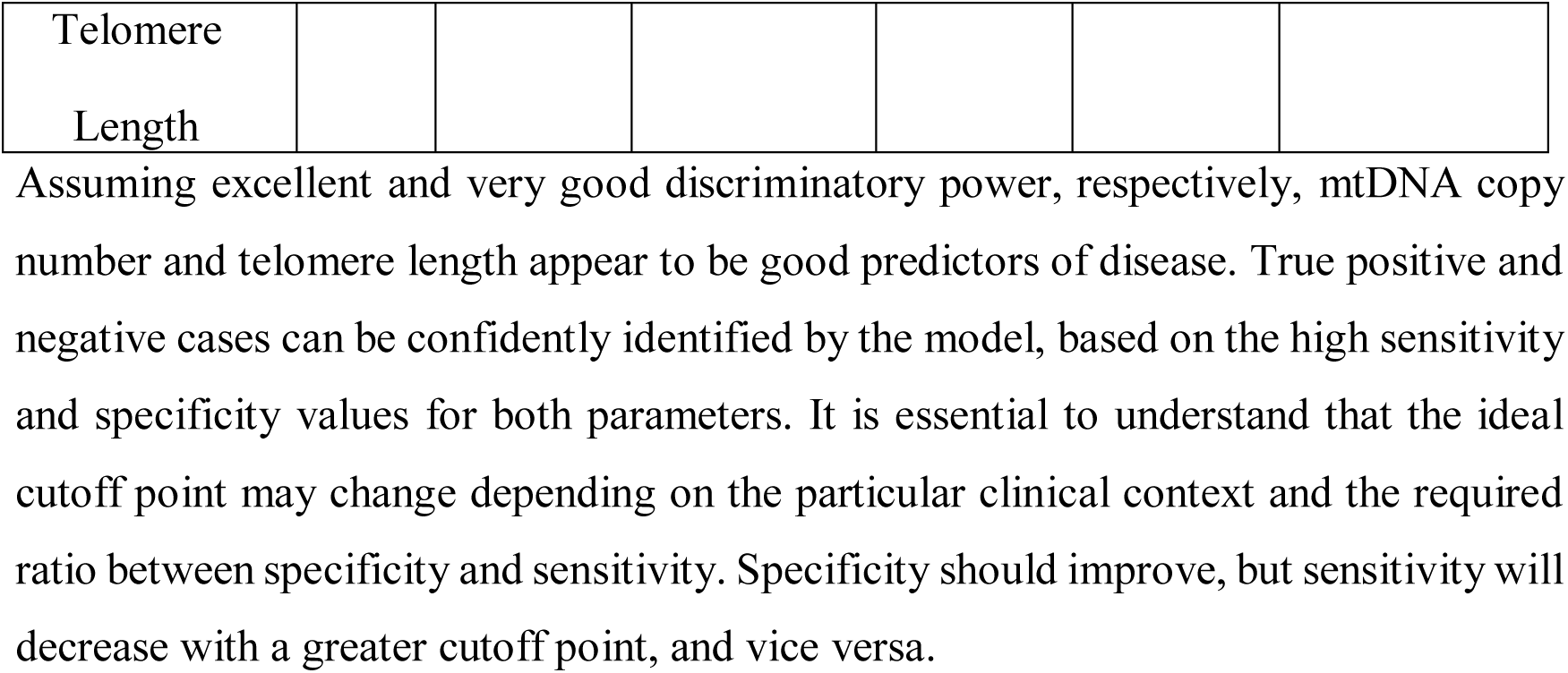
Receiver operating characteristics curve (ROC)

## Discussion

The increased mtDNA copy number in patients suggests that mtDNA replication is dysregulated. The underlying reasons of this instability may be environmental, genetic, or a combination of all three. This could cause mitochondria to create excessive energy, which could lead to toxic reactive oxygen species that would seriously impact the sperm DNA and other components[20]. Elevated sperm mtDNA copy number is associated with damage to sperm and poorer measures of semen quality[21]. Mitochondria, the energy powerhouses of the cell, are essential for sperm health. An increase in mtDNA copy number can impair energy production, leading to decreased sperm motility, viability, and overall fertility[22]. Sperm mitochondrial function, encompassing energy production and apoptosis regulation, is vital during spermatogenesis, while sperm mtDNA copy number is physiologically reduced during sperm maturation, elevated levels are associated with compromised semen quality, as shown in meta-analytic studies [21].

Mitochondria are essential for male fertility, playing a key role in germ cell development and sperm function. While ROS promote sperm capacitation, excessive levels induce oxidative stress, impairing sperm quality and motility. Thus, mitochondrial assessment is essential to understand male infertility [23]. Additionally, mitochondria generate reactive oxygen species (ROS), which can damage sperm DNA and other cellular components if not balanced with antioxidants. This oxidative stress can contribute to infertility[24].

Precise regulation of mtDNA copy number is fundamental to reproductive success; A high mtDNA copy number ensures the proper distribution of mitochondria, necessary for early embryonic development. Conversely, itsdownregulationis essential for optimal sperm function. Furthermore, an increase in mtDNA copy number can trigger apoptosis, or programmed cell death, leading to a decrease in sperm countand affecting overall fertility. Finally, mtDNA is involved in sperm maturation, and anincrease in mtDNA copy number can disrupt this process, leading to abnormal sperm development and reduced fertility[10,23,25].

Male infertility can be caused by a variety of factors, including age, lifestyle, and environmental exposures, which can affect mtDNAcopy number. Measuring mtDNA copy number may prove to be a useful biomarker for identifying men who would benefit from particular treatments and for detecting male infertility[26]. A previous investigation by Song and Lewis (2008) reported similar results to our study. Their research examined mtDNA integrity and copy number in sperm from infertile men and found that individuals with oligoasthenoteratozoospermia (OATS) had a significantly higher mean mtDNA copy number compared to those with normal semen parameters who had asthenozoospermia. Specifically, they found that the mean mtDNA copy numbers was 15.7 in men with normal parameters, 34.3 in asthenozoospermia, 56.7 in oligozoospermia, and 73.7 in OATS [27]. Wu et al. conducted a study investigating the relationship between sperm mtDNA copy number, deletion rate, and infertility. Their findings suggest that high mtDNA copy number in sperm is linked to male infertility, highlighting its potential as a sensitive biomarker to assess male reproductive health [28]. Similar to the observations in our study, Jiang et al. (2017), show that increasing mtDNA copy number can successfully reverse severe disease features resulting from mtDNA mutations, thus confirming the effectiveness of this strategy for treating mitochondrial disorders[29].

Telomere length has long been associated with aging. Telomeres act as protective caps on chromosomes, playing a crucial role in maintaining genomic integrity and cellular function. There is a strong link between aging and infertility in both sexes, with women often becominginfertile at an earlier age. In the last decade, telomeres have gained significant attention due to their role in fertility[30]. Telomeres, repetitive noncoding DNA sequences, play a criticalrole in chromatin integrity. Telomere length is age-dependent in somatic cells, but increases with age in sperm cells [31]. Sperm quality depends on various factors such as sperm count, motility, viability, reactive oxygen species (ROS) level, DNA fragmentation index (DFI), and telomere length. Sperm telomere length is critical during spermatogenesis, fertilization, pronucleus formation, and meiosis, although the exact mechanisms regulating sperm telomere length in male infertility are not fully understood. Sperm telomere length is directly associated with vitality, protamination, and progressive motility, and is negatively associated with DNA fragmentation. The link between sperm telomere length and its impact on male fertility is likely related to increased oxidative stress. Oxidative stress significantly damages hematopoietic stem cells and has been shown to be responsible for dysfunction and aging of somatic and germ cells. Severe oxidative stress is one of the main factors responsible for male infertility and telomere shortening. Telomeres are rich in guanine residues, which are susceptible to oxidative stress, leading to increased sperm DNA damage, thereby reducing sperm quality and leading to infertility [32].

The results of the present study are consistent with those of Margiana et al,. (2024), who showed that telomere length was decreased in the oligospermia group (p < .0001) compared to the healthy group[33]. Similarly, the results are consistent with the study conducted by Dhillon et al. (2024), who assessed that the absolute length of sperm telomere was significantly shorter (p = 0.004) in oligospermic individuals compared to the fertile group, and observed a significant positive correlation between absolute sperm telomere length and sperm parameters[34].

Similarly, Kate et al. (2023) showed that the mean (SE) of Sperm Telomere Length (STL), recorded in infertility cases was significantly shorter than that of the control group; 140.60 (6.66) kb/genome and 239.63 (12.32) kb/genome respectively (p <0.001). A moderate positive correlation was evident between STL in kb/genome and the total sperm count in millions/ml (rho= 0.54, p<0.001), progressive sperm motility (rho= 0.56, p=<0.001) and sperm viability (rho= 0.51 p=0.032) in the infertile group[35]. Moreover, the result of the current study was consistent with another study reported by Torra et al. (2018), who showed that there were strong associations between sperm telomere length and sperm count; i.e., men with longer sperm telomeres tend to have better sperm count than those with shorter telomeres. Thus, sperm telomere length can provide information about male fertility, as a shortened telomere can be a sign of impaired spermatogenesis, which can lead to a low sperm count, errors in chromosome segregation and imbalanced gametes[36].

Furthermore, the results in the current study are consistent with those obtained by Darmishonnejad et al. (2020), who revealed that sperm telomere length were significantly shorter in infertile men than in fertile individuals, and observed significant associations between telomere length with sperm concentration, DNA fragmentation and lipid peroxidation[31]. Furthermore, increased oxidative stress in sperm from infertile men may lead to abnormal chromatin packaging, DNA damage and shorter sperm telomere length. Both mtDNA copy number and STL are indicators of cellular aging and have a comparable impact. Cellular function is compromised by telomere shortening and decreaingmtDNA copy number with cellular aging. Male fertility may be more significantly affected by the combination of short telomeres and decrease mtDNA copy number than by either condition alone[37]. A previous study opposite to the current finding, found a consistent positive correlation between mtDNA copy number and telomere length in adults, suggesting a co-regulatory link between these factors, which may impact age-related and stress-related health problems [38].

The complex relationship between male infertility, telomere length, and mtDNA copy number requires further investigation. This includesinvestigating the fundamental processes that link these variables and identifying possible treatment targets to increase male fertility[39].

Male fertility is greatly impacted by the critical roles that telomeres and mitochondria play in cellular health and function. Several studies have demonstrated that sperm with reduced copies of mitochondrial DNA become less motile and viable, negatively impacting fertility. Telomeres and mtDNA have a reciprocal interaction in which each has a consequence on the other. Telomere shortening, for example, can be accelerated by increased oxidative stress caused by aincrease in mtDNA copies number [23,40].

Therefore, the combination of shortened telomeres and decreasedmtDNA copy number affects mitochondrial function, reducing sperm energy production, causing DNA damage, and ultimately increasing the risk of infertility. Research in this area is essential to create new diagnostic tools to assess sperm quality and identify the fundamental causes of male infertility. It is also essential to create creative approaches to treat male infertility [10,22]

## Conclusion

The results reveal that, compared to healthy controls, patients with different degrees of male infertility have significant variations in mtDNA copy number and telomere length. The high mtDNA copy number of the patient group suggests dysregulationof mtDNA replication. Environmental and genetic variables may be responsible for this dysregulation. The results reveal that male fertility and sperm telomere length have a significant association, with shorter telomere length corresponding to decreased sperm quality and a greater likelihood of infertility. In this case, oxidative stress is essential because it causes DNA damage and telomere shortening. In order to comprehend the causes of infertility and to create new therapeutic approaches, it is essential to study the high mtDNA copy number in infertile Iraqi men as diagnostic and prognostic indicators that may shed light on the underlying causes of male infertility. These findings support reproductive health research and improve men’s health and psychological well-being.

## Data Availability

All relevant data are within the manuscript and its Supporting Information files

## Author Contributions

Mustafa Faeq Kadhim conceived the study, designed the experiments, collected and analyzed the data, and wrote the manuscript. Ali Gargouri contributed to project supervision, presentation of the results and manuscript editing. Farah Thamer Samawi supervised the project, methodology and reviewed the manuscript. Mehdi Elarbi had role in data analysis and interpretation.

## Availability of data

data will be made available upon study completion

## Notes

### Competing Interest Statement

The authors have declared no competing interest.

### Funding Statement

The author(s) received no specific funding for this work.

### Author Declarations

The Ethics Committee of Al-Nahrain University gave ethical approval for this work. Full names of members of "Ethics Committee": - Head of Committee: Prof. Dr. Rebah Najah Algafari Head of environmental biotechnology department. Biotechnology research center Al-Nahrain University -Member of Committee: Assist. Prof. Dr. Zaid Akram Al-Rawi Head of the Quality Department Presidency of Al-Nahrain University - Member of Committee: Assist. Prof. Dr. Omar Abid Kathum Researcher in plant biotechnology dept. Biotechnology research center Al-Nahrain University

## References

[1] Dourou P, Gourounti K, Lykeridou A, Gaitanou K, Petrogiannis N, Sarantaki A. Quality of Life among Couples with a Fertility Related Diagnosis. Clin Pract 2023;13:251–63. 10.3390/clinpract13010023.

[2] Jaiswal A, Baliu-Souza T, Turner K, Nadiminty N, Rambhatla A, Agarwal A, et al. Sperm centriole assessment identifies male factor infertility in couples with unexplained infertility – a pilot study. Eur J Cell Biol 2022;101:151243. 10.1016/j.ejcb.2022.151243.

[3] Legese N, Tura AK, Roba KT, Demeke H. The prevalence of infertility and factors associated with infertility in Ethiopia: Analysis of Ethiopian Demographic and Health Survey (EDHS). PLoS One 2023;18:e0291912. 10.1371/journal.pone.0291912.

[4] Deshpande PS, Gupta AS. Causes and Prevalence of Factors Causing Infertility in a Public Health Facility. J Hum Reprod Sci 2019;12:287–93. 10.4103/jhrs.JHRS_140_18.

[5] Katz-Wise SL, Priess HA, Hyde JS. Gender-role attitudes and behavior across the transition to parenthood. Dev Psychol 2010;46:18–28. 10.1037/a0017820.

[6] Hussam F, Abdulhameed Khudair S, K. Alkhafaje W, S. Alnassar Y, M. Kaoud R, Najm Abed A, et al. A Cross-Sectional Study Regarding Infertility Among Women in Iraq. J Obstet Gynecol Cancer Res 2022;8:47–52. 10.30699/jogcr.8.1.47.

[7] Mohammed T, Burhan S, Ali Hamza N, Kathem A. Ratio of Male to Female Infertility in Baghdad Al-Karkh (2015-2020). Teikyo Med J 2021;44:2197–207.

[8] Rossmann MP, Dubois SM, Agarwal S, Zon LI. Mitochondrial function in development and disease. Dis Model Mech 2021;14. 10.1242/dmm.048912.

[9] Ferreira T, Rodriguez S. Mitochondrial DNA: Inherent Complexities Relevant to Genetic Analyses. Genes (Basel) 2024;15. 10.3390/genes15050617.

[10] Moustakli E, Zikopoulos A, Skentou C, Bouba I, Tsirka G, Stavros S, et al. Sperm Mitochondrial Content and Mitochondrial DNA to Nuclear DNA Ratio Are Associated with Body Mass Index and Progressive Motility. Biomedicines 2023;11. 10.3390/biomedicines11113014.

[11] Filograna R, Mennuni M, Alsina D, Larsson N-G. Mitochondrial DNA copy number in human disease: the more the better? FEBS Lett 2021;595:976–1002. 10.1002/1873-3468.14021.

[12] Maggo S, North LY, Ozuna A, Ostrow D, Grajeda YR, Hakimjavadi H, et al. A method for measuring mitochondrial DNA copy number in pediatric populations. Front Pediatr 2024;12. 10.3389/fped.2024.1401737.

[13] Zhang Y, Ma Y, Bu D, Liu H, Xia C, Zhang Y, et al. Deletion of a 4977-bp Fragment in the Mitochondrial Genome Is Associated with Mitochondrial Disease Severity. PLoS One 2015;10:e0128624. 10.1371/journal.pone.0128624.

[14] Faeqali Jan M, Muneer Al-Khafaji H, Hasan Al-Saadi B, Aneed Al-Saedi MK. Assessment of Interleukin-8 in Bronchial Asthma in Iraq. Arch Razi Inst 2021;76:913–23. 10.22092/ari.2021.355733.1712.

[15] Bertolo A, Capossela S, Fränkl G, Baur M, Pötzel T, Stoyanov J. Oxidative status predicts quality in human mesenchymal stem cells. Stem Cell Res Ther 2017;8:3. 10.1186/s13287-016-0452-7.

[16] Lin J, Smith DL, Esteves K, Drury S. Telomere length measurement by qPCR – Summary of critical factors and recommendations for assay design. Psychoneuroendocrinology 2019;99:271–8. 10.1016/j.psyneuen.2018.10.005.

[17] SPSS. Statistical Package for the Social Sciences 2019:Version 26.

[18] Schmittgen TD, Livak KJ. Analyzing real-time PCR data by the comparative C(T) method. Nat Protoc 2008;3:1101–8. 10.1038/nprot.2008.73.

[19] Riedel G, Rüdrich U, Fekete-Drimusz N, Manns MP, Vondran FWR, Bock M. An extended ΔCT-method facilitating normalisation with multiple reference genes suited for quantitative RT-PCR analyses of human hepatocyte-like cells. PLoS One 2014;9:e93031.

[20] Mengel-From J, Thinggaard M, Dalgård C, Kyvik KO, Christensen K, Christiansen L. Mitochondrial DNA copy number in peripheral blood cells declines with age and is associated with general health among elderly. Hum Genet 2014;133:1149–59. 10.1007/s00439-014-1458-9.

[21] Aisyah CR, Mizuno Y, Masuda M, Iwamoto T, Yamasaki K, Uchida M, et al. Association between Sperm Mitochondrial DNA Copy Number and Concentrations of Urinary Cadmium and Selenium. Biol Trace Elem Res 2024;202:2488–500.

[22] Vahedi Raad M, Firouzabadi AM, Tofighi Niaki M, Henkel R, Fesahat F. The impact of mitochondrial impairments on sperm function and male fertility: a systematic review. Reprod Biol Endocrinol 2024;22:83. 10.1186/s12958-024-01252-4.

[23] Costa J, Braga PC, Rebelo I, Oliveira PF, Alves MG. Mitochondria Quality Control and Male Fertility. Biology (Basel) 2023;12. 10.3390/biology12060827.

[24] Agarwal A, Virk G, Ong C, du Plessis SS. Effect of oxidative stress on male reproduction. World J Mens Health 2014;32:1–17. 10.5534/wjmh.2014.32.1.1.

[25] Wai T, Ao A, Zhang X, Cyr D, Dufort D, Shoubridge EA. The role of mitochondrial DNA copy number in mammalian fertility. Biol Reprod 2010;83:52–62. 10.1095/biolreprod.109.080887.

[26] Mai Z, Yang D, Wang D, Zhang J, Zhou Q, Han B, et al. A narrative review of mitochondrial dysfunction and male infertility. Transl Androl Urol Vol 13, No 9 (September 30, 2024) Transl Androl Urol 2024.

[27] Song GJ, Lewis V. Mitochondrial DNA integrity and copy number in sperm from infertile men. Fertil Steril 2008;90:2238–44.

[28] Wu H, Whitcomb BW, Huffman A, Brandon N, Labrie S, Tougias E, et al. Associations of sperm mitochondrial DNA copy number and deletion rate with fertilization and embryo development in a clinical setting. Hum Reprod 2019;34:163–70.

[29] Jiang M, Kauppila TES, Motori E, Li X, Atanassov I, Folz-Donahue K, et al. Increased Total mtDNA Copy Number Cures Male Infertility Despite Unaltered mtDNA Mutation Load. Cell Metab 2017;26:429–436.e4. 10.1016/j.cmet.2017.07.003.

[30] Vasilopoulos E, Fragkiadaki P, Kalliora C, Fragou D, Docea AO, Vakonaki E, et al. The association of female and male infertility with telomere length (Review). Int J Mol Med 2019;44:375–89. 10.3892/ijmm.2019.4225.

[31] Darmishonnejad Z, Tavalaee M, Zarei Kheirabadi M, Zohrabi D, Nasr-Esfahani MH. Relationship between sperm telomere length and sperm quality in infertile men. Andrologia 2020;52:e13546. 10.1111/and.13546.

[32] Amir S, Vakonaki E, Tsiminikaki K, Tzatzarakis N. M, Michopoulou V, Flamourakis M, et al. Sperm telomere length: Diagnostic and prognostic biomarker in male infertility (Review). World Acad Sci J 2019;1:259–63. 10.3892/wasj.2020.31.

[33] Margiana R, Gupta R, Al-Jewari W, Hjazi A, Alsaab H, Singh R, et al. Evaluation of telomere length, reactive oxygen species, and apoptosis in spermatozoa of patients with oligospermia. Cell Biochem Funct 2024;42:e3935. 10.1002/cbf.3935.

[34] Dhillon VS, Shahid M, Deo P, Fenech M. Reduced SIRT1 and SIRT3 and Lower Antioxidant Capacity of Seminal Plasma Is Associated with Shorter Sperm Telomere Length in Oligospermic Men. Int J Mol Sci 2024;25. 10.3390/ijms25020718.

[35] Rajesh K, Vatsalaswamy P, Manvikar P. The association of male infertility with telomere length: A case control study. Indian J Clin Anat Physiol 2021;8:325–32. 10.18231/j.ijcap.2021.070.

[36] Torra-Massana M, Barragán M, Bellu E, Oliva R, Rodríguez A, Vassena R. Sperm telomere length in donor samples is not related to ICSI outcome. J Assist Reprod Genet 2018;35:649–57. 10.1007/s10815-017-1104-2.

[37] Yan X, Yang P, Li Y, Liu T, Zha Y, Wang T, et al. New insights from bidirectional Mendelian randomization: causal relationships between telomere length and mitochondrial DNA copy number in aging biomarkers. Aging (Albany NY) 2024;16:7387–404. 10.18632/aging.205765.

[38] Tyrka AR, Carpenter LL, Kao H-T, Porton B, Philip NS, Ridout SJ, et al. Association of telomere length and mitochondrial DNA copy number in a community sample of healthy adults. Exp Gerontol 2015;66:17–20. 10.1016/j.exger.2015.04.002.

[39] Moustakli E, Zikopoulos A, Skentou C, Dafopoulos S, Stavros S, Dafopoulos K, et al. Association of Obesity with Telomere Length in Human Sperm. J Clin Med 2024;13. 10.3390/jcm13072150.

[40] Park Y-J, Pang M-G. Mitochondrial Functionality in Male Fertility: From Spermatogenesis to Fertilization. Antioxidants (Basel, Switzerland) 2021;10. 10.3390/antiox10010098.

